# JAK/STAT Signaling Predominates in Human and Murine Fungal Post-infectious Inflammatory Response Syndrome

**DOI:** 10.1101/2024.01.18.24301483

**Authors:** Jessica C. Hargarten, Kenneth Ssebambulidde, Seher H. Anjum, Malcolm J. Vaughan, Jintao Xu, Brian Song, Anutosh Ganguly, Yoon-dong Park, Terri Scott, Dima A. Hammoud, Michal A. Olszewski, Peter R. Williamson

**Affiliations:** Laboratory of Clinical Immunology and Microbiology (LCIM), Division of Intramural Research (DIR), National Institute of Allergy and Infectious Diseases (NIAID), National Institutes of Health (NIH), Bethesda, MD, USA; Division of Pulmonary and Critical Care Medicine, Department of Internal Medicine, University of Michigan Health System, Ann Arbor, MI, USA; Research Service, Ann Arbor VA Healthcare System, Department of Veterans Affairs Health System, Ann Arbor, MI, USA; Infectious Diseases Institute, College of Health Sciences, Makerere University, Kampala, Uganda; Center for Infectious Disease Imaging (CIDI), Radiology and Imaging Sciences, Clinical Center, National Institutes of Health, Bethesda, Maryland, USA

## Abstract

Post-infection inflammatory syndromes have been increasingly recognized as a cause of host damage in a variety of infectious diseases including tuberculosis, bacterial meningitis, and COVID-19. Recently, a post-infectious inflammatory response syndrome (PIIRS) was described in non-HIV-infected cryptococcal fungal meningoencephalitis (CM) as a major cause of mortality. Inflammatory syndromes are particularly severe in neurological infections due to the skull’s rigid structure which limits unchecked tissue expansion from inflammatory-induced edema. In the present studies, neurologic transcriptional pathway analysis utilizing a murine PIIRS model demonstrated a predominance of Janus kinase/signal transducer and activator of transcription (JAK/STAT) activation. JAK/STAT inhibitor treatment resulted in improvements in CNS damage markers, reductions in intrathecal CD44^hi^CD62^lo^ CD4^+^ effector CD4^+^ T-cells and MHC II^+^ inflammatory myeloid cells, and weight gains in mice, the latter after treatment with antifungals. Based on these data, pathway-driven steroid-sparing human treatment for steroid-refractory PIIRS was initiated using short courses of the JAK/STAT inhibitor ruxolitinib. These were well tolerated and reduced activated HLA-DR^+^ CD4^+^ and CD8^+^ cells and inflammatory monocytes as well as improved brain imaging. Together, these findings support the role of JAK/STAT in PIIRS as well as further study of JAK/STAT inhibitors as potential adjunctive therapy for PIRS and other neural inflammatory syndromes.

## Introduction

Post-infection inflammatory complications have recently gained considerable attention, especially for bacterial and viral meningitides and post-acute sequelae of SARS-CoV2 infection (PASC) (1, 2). Neurologic inflammatory syndromes have also been described in infections with the neurotropic fungus *Cryptococcus* with both HIV-related immune reconstitution syndromes (IRIS) and non-HIV-related inflammatory response syndromes (PIIRS), currently being the largest cause of non-viral meningitis in the US (3). In these latter syndromes, the skull’s rigid structure limits unchecked tissue expansion from inflammatory-induced edema. The compression can result in a neurological deterioration or even fatal uncal herniation due to rising intracranial pressure (4). Distinctively, PIIRS manifests without clear immune reconstitution and results in clinical deterioration likely from antigen release during anti-fungal therapy despite concordant negative cerebral spinal fluid cultures (5). Cohort studies have found pulse-corticosteroid taper therapy (PCT) effective against this inflammation (6), leading to prompt clinical improvement in all treated patients. Symptom alleviation has been linked to reductions in multiple CSF inflammatory markers, including activated HLA-DR^+^ CD4^+^ and CD8^+^ T cells, inflammatory monocytes, glucose, protein, and levels of soluble CD25 (IL-2R) and interleukin-6 (IL-6) (6). Most patients, without prior health conditions and diagnosed with PIIRS, have been able to reduce their dependency on steroids after a year post-treatment initiation (3). This reduction rate is notably slower compared to conditions like cIRIS and TB meningitis (7), leading to potentially increased rates of steroid-related complications (8). Progress in developing effective, rationally targeted therapies for PIIRS has been limited by our lack of mechanistic understanding of the underlying key inflammatory pathways in PIIRS.

To uncover molecular pathways involved in the pathogenesis of PIIRS, we conducted neurological transcriptional pathway analysis of NanoString-identified transcripts from brains utilizing a recently-developed mouse model of PIIRS (9). These studies identified a predominance of Signal Transducer and Activator of Transcription 1 (STAT1) and STAT3-associated downstream genes. Since Janus kinase (JAK)-dependent phosphorylation is known to activate both pathways, western blots were then used to demonstrate increased phospho-STAT1 and STAT3 as a measure of activation concurrent with the onset of PIIRS. In addition, treatment of mice with the JAK/STAT pathway inhibitor ruxolitinib during the onset of brain inflammation led to reductions in JAK/STAT transcriptional targets in brains as well as p-STAT1/STAT3 on western blots, brain edema, and brain-infiltrating T cells and monocytes, but clinical improvement was complicated by increases in brain fungal loads. Treatment with the antifungal amphotericin B, as is typical in patients, resulted in equivalence between the untreated and ruxolitinib-treated mice in brain fungal loads, which then resulted in weight gains in the ruxolitinib-treated mice. These encouraging pre-clinical studies led to the use of short courses of ruxolitinib in four patients with refractory PIIRS who exhibited continued inflammation despite corticosteroid therapy and one patient who was not a good candidate for corticosteroids. In these patients, ruxolitinib was well tolerated with reduced activated HLA-DR^+^ CD4^+^ and CD8^+^ cells and inflammatory monocytes as well as soluble markers IL-6 and sCD25. Because of significant clinical benefit in one patient who had not been able to reduce corticosteroids, we continued ruxolitinib for 1 year with apparent clinical benefit, enabling tapering of corticosteroid therapy. In summary, these findings support a prominent role for the JAK/STAT pathway in PIIRS and provide proof-of-concept for further study of ruxolitinib as a novel potential adjunctive therapy for clinical trials in PIIRS.

## Results

### PIIRS is associated with significant upregulated inflammatory parameters and evidence of increased signaling of the JAK/STAT pathways

To identify candidate inflammatory pathways involved with PIIRS, we utilized a recently-described cryptococcal PIIRS mouse model ((9), Fig. 1a). Brain homogenates at the peak of inflammation (21 days post-infection (dpi)) underwent NanoString Neuroinflammation transcriptional analysis (10–13). Ingenuity Pathway Analysis was performed to rank the top transcriptional regulators of PIIRS-associated neuroinflammation and identified Janus kinase (JAK)/STAT signaling, particularly STAT1 and STAT3 as the top 5 regulators and highly significant (Fig. 1b). To determine whether inhibition of the JAK signaling would attenuate PIIRS disease severity, *Cn* infected mice were randomized to either ruxolitinib (1 g/kg daily) or vehicle alone (Nutragel) according to a previously-described ruxolitinib therapeutic model (14). Drug inhibition was utilized rather than genetic suppression using knockout mice because available mouse strains would not allow short periods of JAK/STAT pathway inhibition that would reduce unrestrained fungal proliferation and rapid mortality. Following ruxolitinib treatment, the JAK/STAT transcriptional signature (Fig. 1b) and STAT1 phosphorylation (Fig. 1c) reduced within the brain of mice. Even with short periods of JAK/STAT inhibition, ruxolitinib treatment led to a reduction in cerebral edema as measured by brain weight (Fig. 1d), but increased fungal burdens (Fig. 1e) resulting in weight loss between 10-14 dpi (Fig. 1f). Consistent with a systemic effect, spleen size and weights (Fig. 1g, h) and splenocyte numbers (Fig. 1i) also showed reductions after treatment. However, addition of the antifungal agent amphotericin B (AmB; Fig. 1j), as performed in clinical care, reduced and normalized brain fungal loads between the treatment groups (Fig. 1k), leading to dramatic weight gains in ruxolitinib-treated mice (Fig. 1l), but without reductions in overall brain weights (Fig. 1m). These data demonstrate the predominance of JAK/STAT pathway activation during PIIRS with improvements after selective JAK/STAT pathway inhibition.

**Fig 1.**
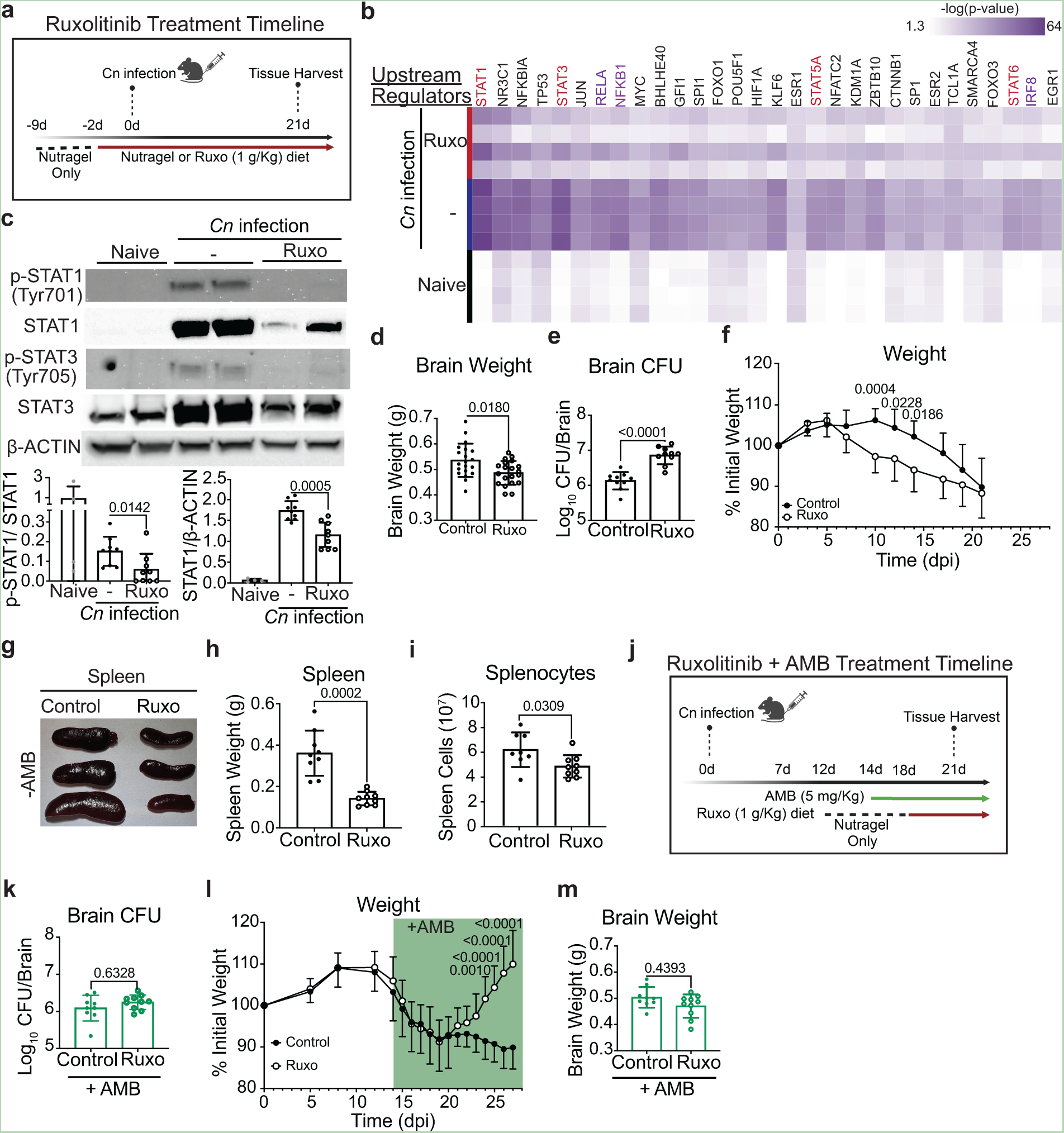
JAK inhibition with daily ruxolitinib treatment in a mouse model of CM-PIIRS attenuates signs of disease and systemic inflammation. a Graphical schema for mouse infection with 10^6^ *Cryptococcus neoformans* 52D (iv) and daily treatment with ruxolitinib (1 g/kg Nutragel diet). At 21 days post-infection (dpi), brain and spleen were harvested. b Heatmap showed top 30 upstream regulators identified using Ingenuity Pathway Analysis (IPA) of NanoString Neuroinflammation Panel gene expression data from whole brain compared to naïve mice (n=4 mice per group). Red regulators=JAK/STAT pathway specific, purple regulators=downstream of JAK/STAT pathway signaling. c Brain homogenates were harvested on 21 dpi for immunoblot analysis of phospho-STAT1 (Tyr701), total STAT1, phospho-STAT3 (Tyr705), total STAT3, and β-actin as loading control. Top, representative protein immunoblot images; bottom, quantification of protein immunoblot data (n=4-8 mice per group; two experiments). d-h Brain weight (d; 21 dpi), brain fungal burden (e; 21 dpi), mouse weights (f; throughout infection), spleen size (g; 21 dpi), spleen weight (h; 21 dpi), and numbers of splenocytes were enumerated (i; 21 dpi). j Graphical schema for mouse infection with 10^6^ *Cryptococcus neoformans* 52D (iv) and delayed amphotericin B (5 mg/kg ip) and ruxolitinib (1 g/kg Nutragel diet) treatment. k-m Brain fungal burden (k; 21 dpi), mouse weights (l; throughout infection), and brain weight (m; 21 dpi) were enumerated as described in . Error bars indicate s.d., n = 8-20 mice per time point. Student’s t-test, p-values indicated above each comparison.

### JAK/STAT inhibition protects mice from neuronal cell apoptotic death and damage to neurotransmission proteins in a mouse model of PIIRS

In order to characterize the neuropathology resulting from excessive JAK/STAT activation in PIIRS, we measured cryptococcal lesions size within brains during onset of inflammation and after inhibition with ruxolitinib. All infected mice developed “Swiss cheese”-like lesions due to *C. neoformans* pseudocyst formation (Fig. 2a). Intriguingly, ruxolitinib-treated mice displayed lesions that were approximately 50% smaller in diameter compared to the other infected groups, either not treated with antifungals or treated with AmB (Fig. 2b). Persistence of the protective effect of ruxolitinib after reduction and normalization of fungal load with AmB suggests the presence of damaging residual inflammation even after fungal load reductions from antifungal therapy as is present in human PIIRS after anti-fungal therapy (5). To determine how treatments modulated the development of neuronal injury and how they can prevent the *C. neoformans*/inflammation-induced neurological damage in CM-PIIRS, we assessed synaptotagmin-7 (Syt7) by immunofluorescence (IF), a protein integral to neurotransmission expression at the perimeter of cryptococcal lesions (15). Our IF data revealed a stark reduction of Syt7 expression in neuronal tissue (visualized by β–III tubulin staining) at the perimeter of the cryptococcal pseudocysts compared to the background expression of Syt7 in similar locations in brains of naïve mice (Fig. 2c, d). Treatment with ruxolitinib but not AmB alone, led to a partial recovery of Syt7 expression. However, the combined treatment of ruxolitinib and AmB fully restored Syt7 expression levels, suggesting a synergistic, protective effect and the full restoration of neuronal health when both drugs were used. Our CM studies using this model previously demonstrated neuronal apoptosis at the perimeter of the cryptococcal micro cysts linked to pathological inflammation (15, 16). To evaluate the effect of the treatments in preventing neuronal apoptosis we also assessed cleaved caspase-3 within β-III tubulin positive neurons (Fig. 2e). The β-III tubulin positive neuronal bundles were negative for cleaved caspase-3 signal in naïve mouse brains, however, in the CM-PIIRS mice, there was a significant presence of cleaved caspase-3 signals, clearly localized with neurons but not CD45^+^ immune cells. Importantly, treatment with ruxolitinib alone or in conjunction with AmB led to a minimal presence of cleaved caspase-3 signals within neurons (Fig. 2f). Inflammatory processes rather than fungal burden thus appeared to be the major driver of lesion enlargement. Indeed, lesions contained significant numbers of CD45 immune cells per lesion, which were reduced after ruxolitinib treatment (Fig. 2g). In conclusion, our findings suggest the role of damaging JAK/STAT inflammation in murine PIIRS and inhibition with ruxolitinib offers protection from neuronal apoptotic cell death and preserves neuronal connectivity proteins.

**Fig 2.**
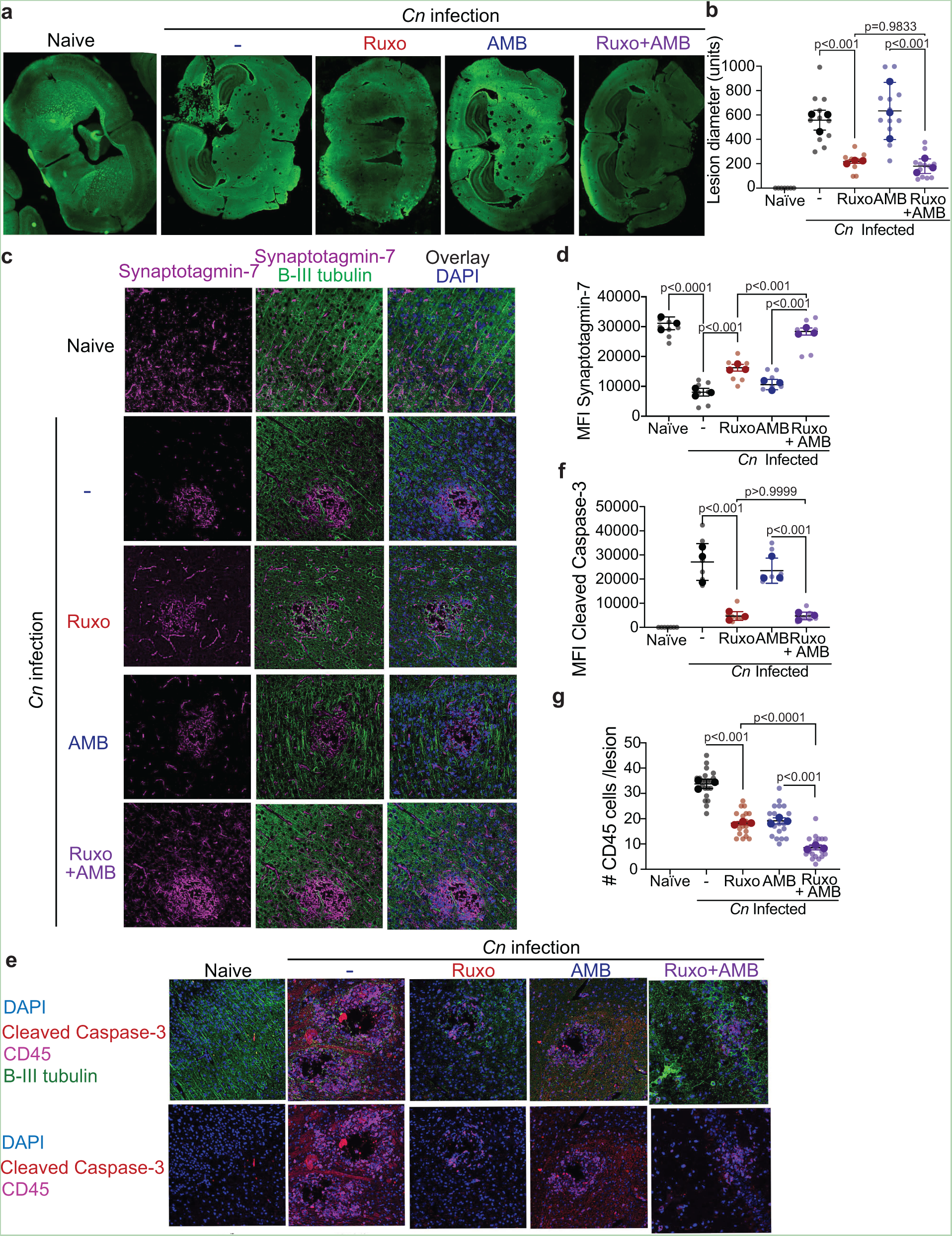
JAK/STAT predominant inflammation contributes to neuronal cell apoptotic death and damage to neurotransmission proteins in a mouse model of CM-PIIRS with protection after ruxolitinib treatment. **a** Immunohistochemistry (IHC) of the brain sections stained with antibodies to beta III tubulin (green) was used to assess the cryptococcal lesions’ size within the brains. **b** Summary statistics of brain lesion sizes from Figure 4a. **c** IHC of the brain sections stained with antibodies to beta III tubulin (green), synaptotagmin-7 (red), and DAPI (blue). The bar graphs on the right show the mean fluorescent intensity (MFI) quantitation of **d** synaptotagmin-7. **e** IHC of brain section stained with antibodies to beta III tubulin (green), cleaved caspase-3 (red), CD45 (pink), and DAPI (blue) with summary statistics of **f** cleaved capspase-3 and **g** CD45^+^ cells/lesion. magnification, 200×. Data shown is representative of two independent experiments (n = 3 mice per group) =/- SEM, and at least 10 fields were examined for each sample. Statistical significance was determined using the linear regression models fit via generalized estimating equations (GEE) with p-values indicated over each comparison.

### JAK inhibition with ruxolitib treatment reduced brain accumulation of activated T cells and inflammatory monocytes in a mouse PIIRS model

Further study sought to characterize key JAK/STAT-related inflammatory cell types involved in PIIRS by assessing for inflammatory changes in the brain. More detailed immunological studies were limited to that without AmB treatment because: 1) AmB has been shown to result in confounding immunological effects of its own, such as promoting innate immune activation, inflammation, and toxicity (17, 18). 2) AmB antifungal activity differs with fungal strains (19), and 3) patients are typically no longer on AmB during subsequent onset and treatment of PIIRS (20). We first assessed global changes in predicted cell type abundance in whole brain homogenates at 21 dpi based on NanoString Neuroinflammation gene expression data and cell profiling analysis (21). Compared to the brain of naïve mice, PIIRS mice show increased abundance of all immune cell types profiled with a decreased abundance of CNS cells (astrocytes, oligodendrocytes, and neurons) (Fig. 3a). Ruxolitinib treatment reversed immune cell accumulation and increased abundance of astrocytes and neurons. To further analyze inflammatory immune populations in the brain at 21dpi, brain infiltrating lymphocytes were isolated and flow cytometry performed which demonstrated the presence of CD45^hi^ lymphocytes that were reduced after treatment with ruxolitinib (Fig. 3b, c). Reductions in brain CD3^+^ T-cells (Fig. 3d,e), CD4^+^ T-cells (Fig. 3f), and CD44^hi^CD62L^lo^ effector CD4 T-cells (Fig. 3g) were also observed after ruxolitinib treatment. Brain inflammatory myeloid populations were also reduced (Fig. 3h-m) after ruxolitinib treatment, specifically inflammatory MHC II^+^ myeloid cells (Fig 3h-j) and activated microglia dually expressing Ly6C^+^ and MHC II^+^ (Fig. 3k-m). These data suggest a role for an orally administered JAK/STAT pathway inhibitor ruxolitinib in limiting recruitment of inflammatory cells to the brain in a murine PIIRS model.

**Fig 3.**
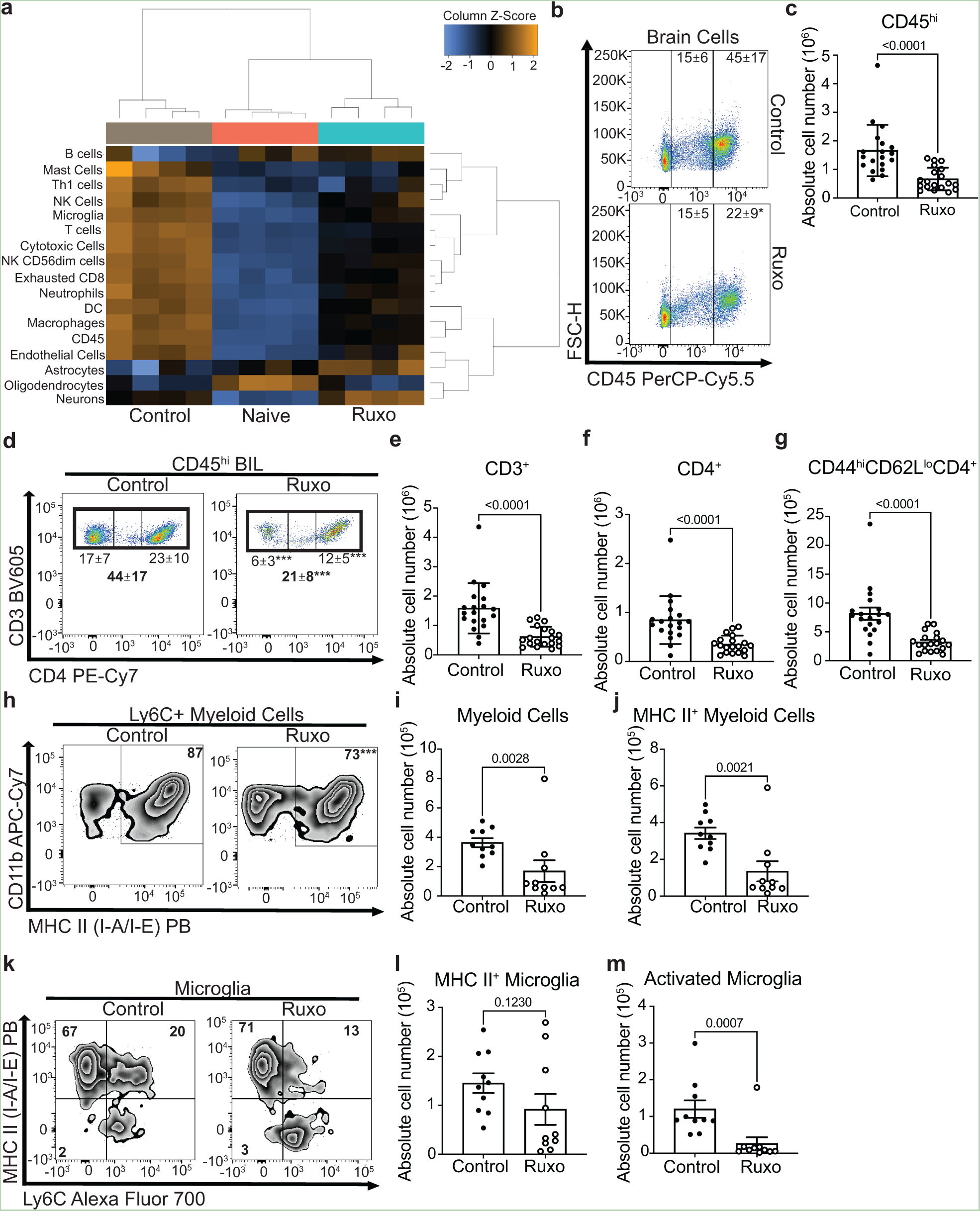
JAK inhibition with daily ruxolitinib treatment in a mouse model of CM-PIIRS reduces accumulation of activated T cells and inflammatory myeloid cells. **a** At 21 dpi, predicted brain cell type abundance was analyzed using NanoString Neuroinflammation Panel gene expression data and cell type abundance profiling analysis. (n=4 mice per group). **b-m** Brains were harvested, brain infiltrating lymphocytes (BIL) isolated, and flow cytometry performed to monitor changes to brain mononuclear cell populations. **b** Live cells were gated and separated based on CD45 expression and flow plots show the percentage of CD45^hi^ and CD45^int^ cells out of the total live brain mononuclear cells and **c** summary statistics show the absolute number of CD45^hi^ brain cells. **d** Representative flow plot and proportion of brain cells that are CD3^+^ and CD4^+^ T cells. **e-g** Number of CD3 T cells (**e**), CD4 T cells (**f**), CD44^hi^CD62L^lo^ effector CD4 T cells **g** from the brain. **h** Representative flow plot and proportion of Ly6C^+^ myeloid cells expressing MHC-II. **i-j** Summary statistics of absolute number of Ly6C^+^ myeloid cells (**i**) and MHC-II+ inflammatory myeloid cells (**j**) in the brain. **k** Representative flow plot and proportion of microglia expressing MHC-II and Ly6C. **l-m** Number of MHC-II^+^ microglia (**l**) and MHC-II^+^/Ly6C^+^ activated microglia (**m**) in the brain. Data shown are the mean ± SD with 10-20 mice per time point and statistical significance was determined by unpaired Student’s t-test.

### Principal component analysis of murine PIIRS brain inflammation demonstrates JAK/STAT-dependent activation of adaptive markers of immunity, cytokine signaling, and cell cytotoxicity damaging pathways

To help identify molecular processes that accompany brain pathology during CM-PIIRS and limited by ruxolitinib treatment, we measured changes in RNA transcripts in the brain on 21 dpi using a NanoString neuroinflammation multiplex panel, which measures the expression of 757 genes involved in neuro-immune processes and interactions. Principal component analysis of normalized gene transcript read counts consistently showed that the naïve mice, untreated/infected, and ruxolitinib-treated/infected mice formed separate clusters by principal components 1 and 2 (Fig. 4a). The top 20 differentially expressed genes (DEG) in the brain of ruxolitinib-treated/infected mice compared to untreated/infected mice were found to be related to cytotoxic lymphocyte granzyme-mediated programmed cell death signaling pathway (18/20 top genes), T cell activation (6/20 top genes), complement pathway activation (14/20 top genes), and cytokine production (4/20 top genes) (Table 1). 6 of these top differentially expressed genes (Lilrb4a, Ptprc, Pros1, C3, Dock2, Fcgr2b) were also found to be downregulated in the brain of naïve mice compared to untreated/infected mice (Table 2). Interestingly, untreated/infected mice showed elevated pathway scores for all neuroinflammation pathways analyzed compared to naïve and ruxolitinib-treated/infected mice (Fig. 4b), particularly adaptive immune responses (Fig. 4c (top)) and cytokine responses (Fig. 4c (bottom)), with pathway scores for ruxolitinib-treated mice uniformly falling between that of naïve and untreated/infected mice, particularly for genes known to be regulated by JAK/STAT pathway signaling such as the cytokines Cxcl9, Cxcl10, Ccl5, and Ccl7 (Fig. 4c). Notably, these transcriptional signatures are echoed in the canonical pathways identified through IPA analysis (Fig. 4d). This indicates that ruxolitinib treatment ameliorates PIIRS-mediated neuroinflammation by driving down-regulation of transcriptional signatures important for adaptive immune activation, cytokine signaling, and cytotoxicity.

**Fig 4.**
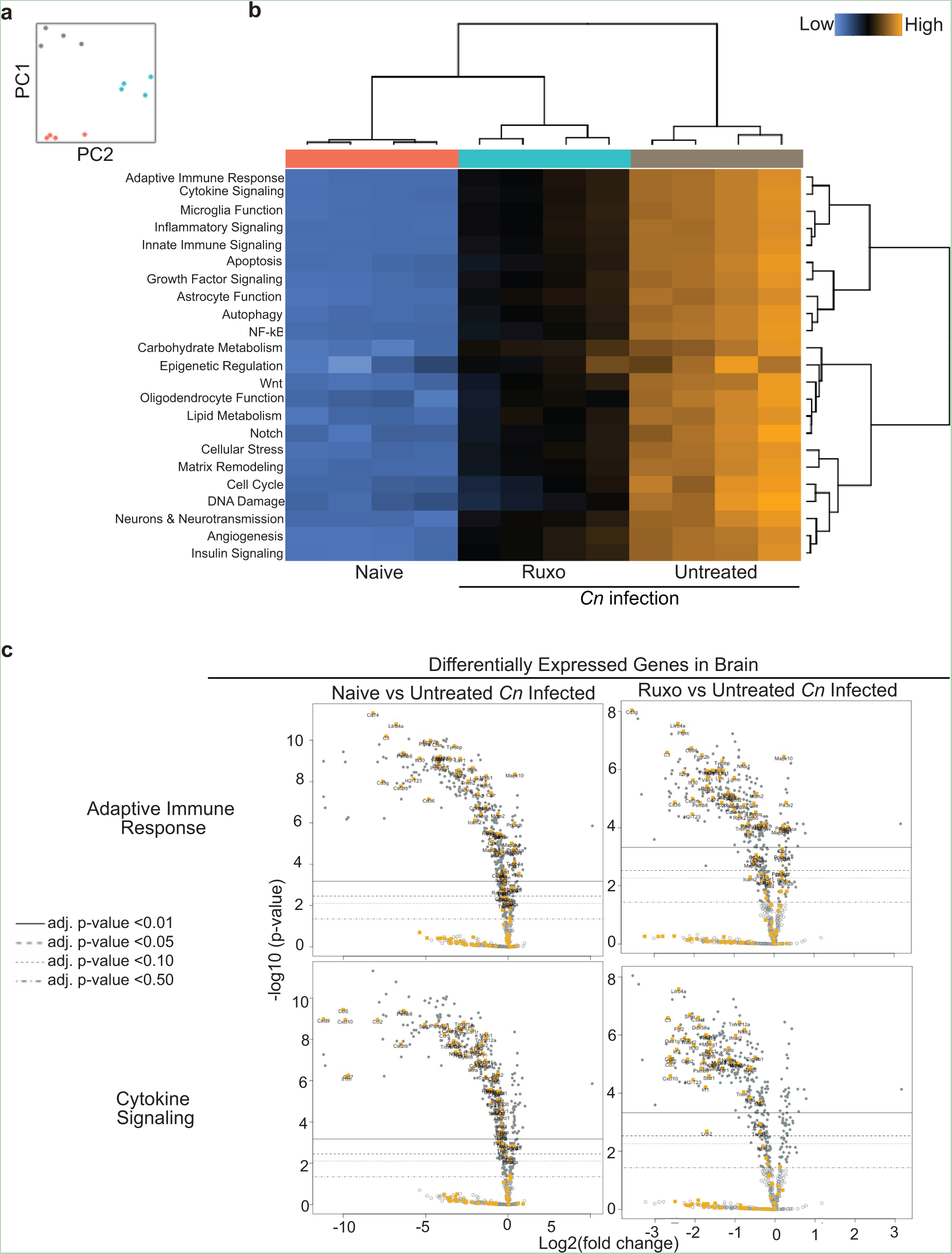
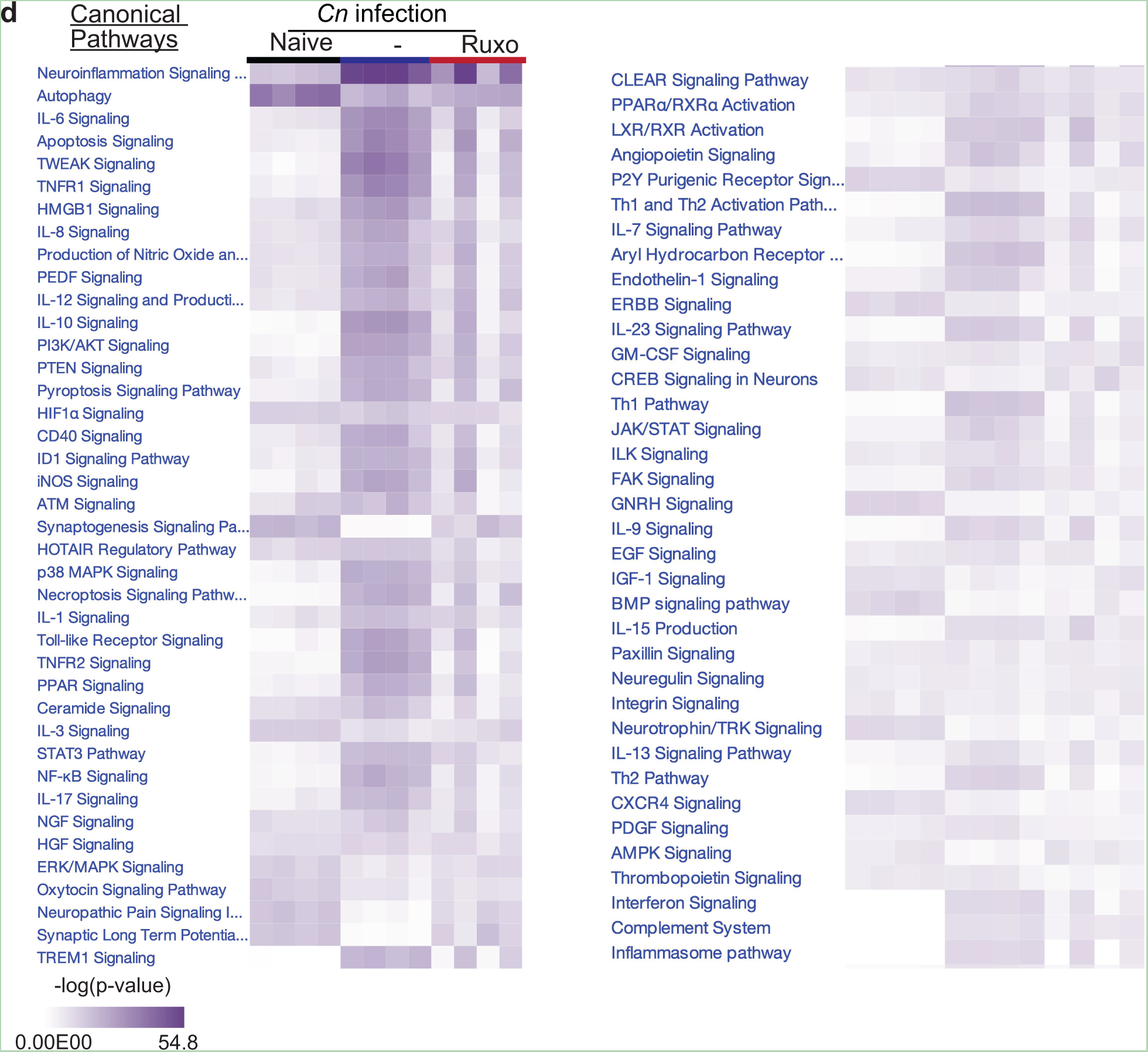
Ruxolitinib ameliorates dysregulated and neuroinflammatory gene expression during murine CM-PIIRS. Gene transcripts of brain homogenates from naïve, untreated/infected, and ruxolitinib-treated/infected mice at 21 dpi were analyzed using a NanoString multiplex neuroinflammation panel. **a** Principal-component analysis (PCA) of normalized read counts of the total of 757 transcripts associated with neuroinflammation. Untreated/Infected and ruxolitinib-treated/infected mice formed separate clusters with principal components 1 and 2. Data from NanoString shown are from one experiment with n=4 mice per group. **b** Ruxolitinib treatment ameliorates all neuroinflammation pathway scores altered by CM-PIIRS in mice. In particular, adaptive immune response and cytokine signaling. **c** Volcano plot of NanoString gene expression analysis comparing brain homogenates from naive or ruxolitinib-treated/infected mice versus untreated/infected mice (N=4 mice per group). Highlighted in orange are differentially expressed genes (DEGs; Log2 fold change (FC)<-1 and adjusted p-value<0.01) belonging to the adaptive immune response pathway (top) or cytokine signaling pathway (bottom). **d** Pathway analysis using DEG between untreated/infected mice and other groups was performed using IPA.

**Table 1.**
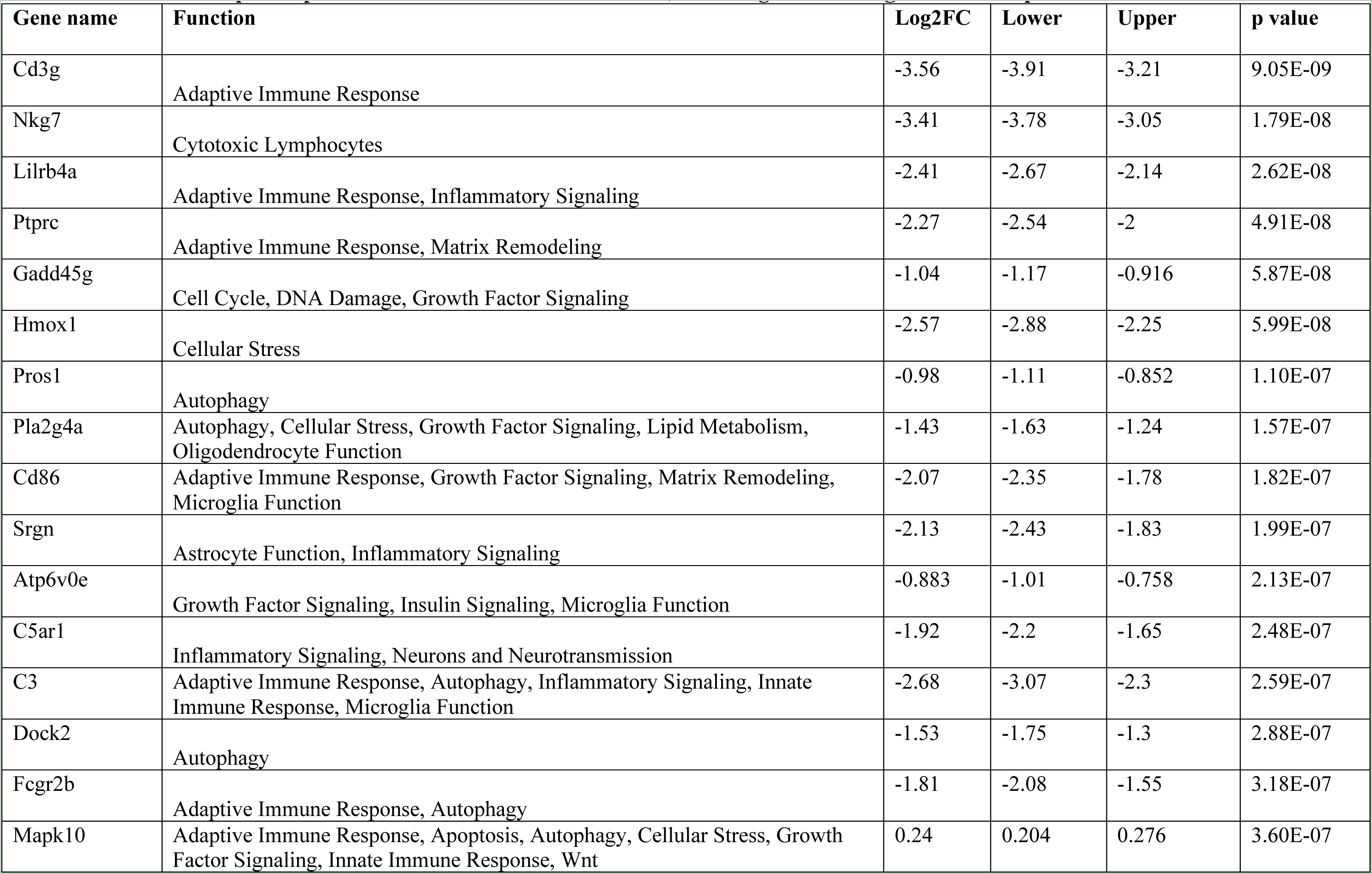
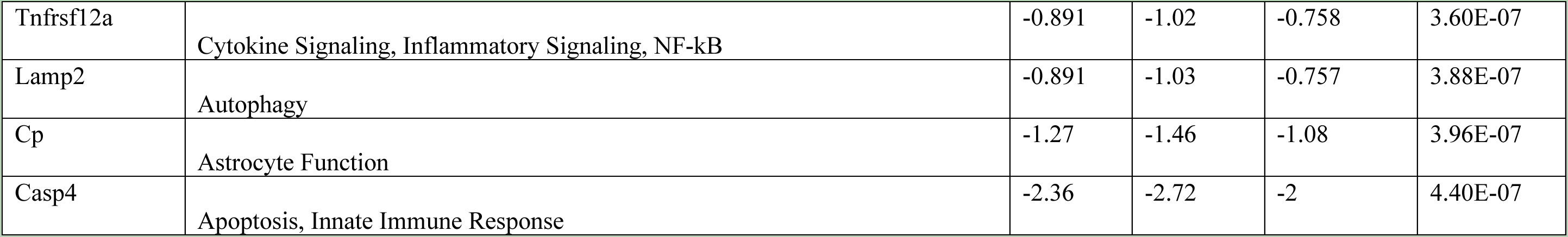
Top 20 differentially expressed genes identified by NanoString analyses in brain homogenates from mice infected and treated with ruxolitinib at 21 dpi compared to infected untreated controls, with Log2 fold-change > 1.15 and p<0.05.

**Table 2.**
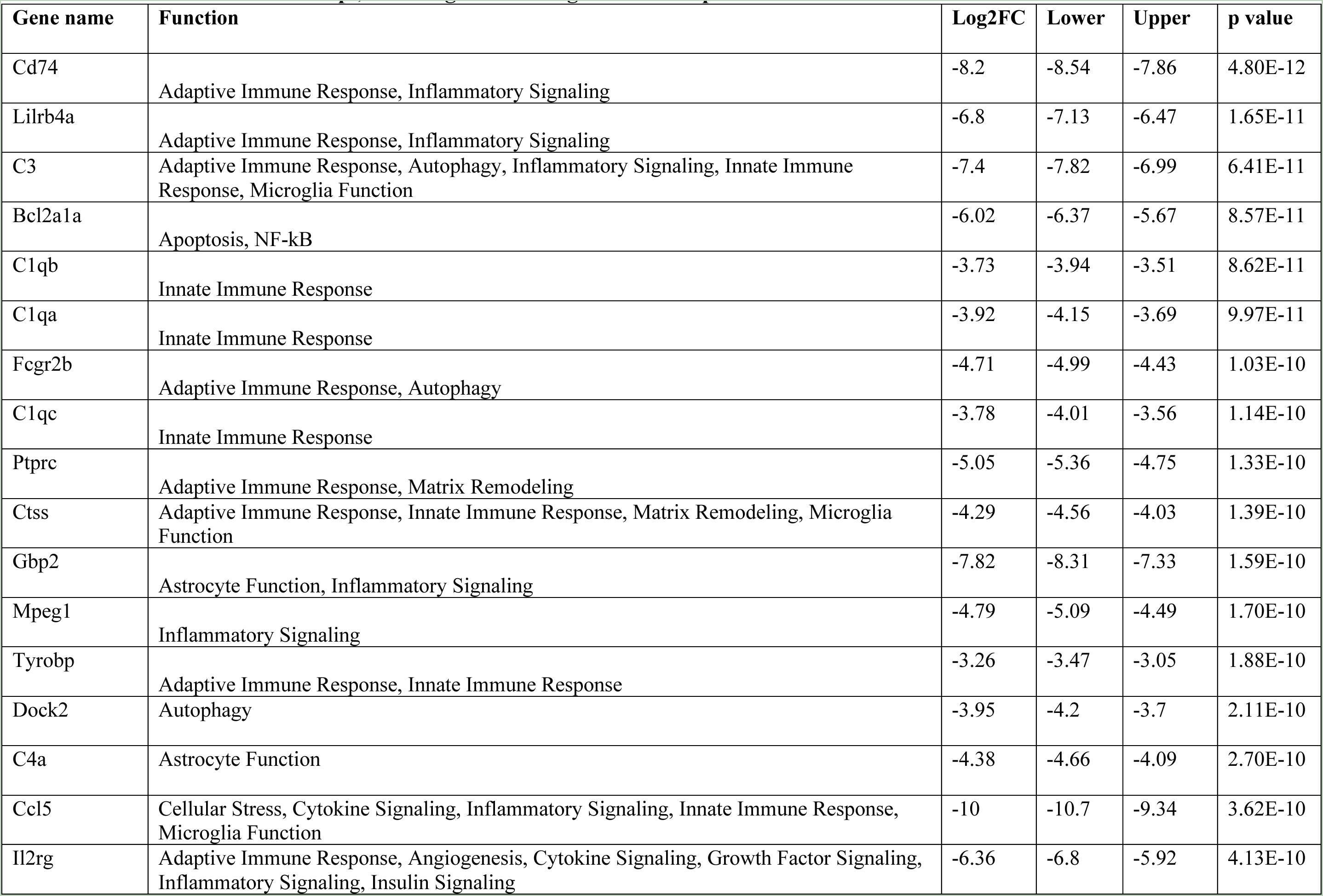
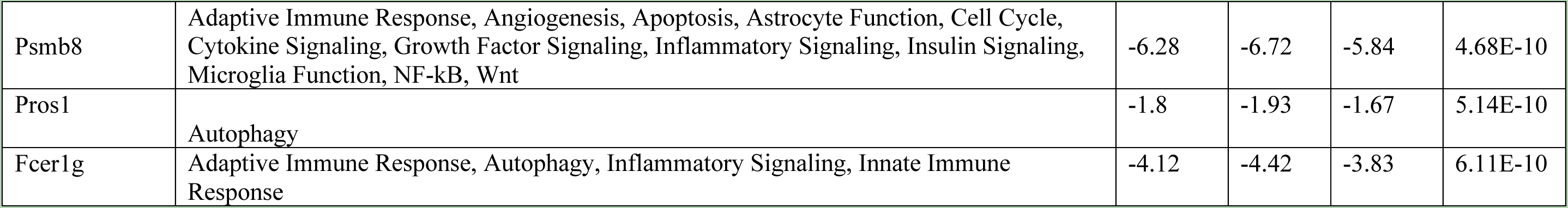
Top 20 differentially expressed genes identified by NanoString analyses in brain homogenates from Naïve mice compared to infected untreated controls at 21 dpi, with Log2 fold-change > 1.15 and p<0.05.

### JAK/STAT pathway inhibition with ruxolitinib reduces CSF inflammatory markers in patients with cryptococcal meningoencephalitis complicated by PIIRS

Based on the above pre-clinical data demonstrating efficacy of ruxolitinib in a mouse model, we undertook a study to see if ruxolitinib in FDA-approved dosages could reduce inflammation in steroid-refractory PIIRS. Five patients diagnosed with cryptococcal meningoencephalitis presented to the NIH Clinical Center under an approved protocol and four were treated with pulse corticosteroid therapy as described (6) but continued to be symptomatic from PIIRS after 1-6 months. Due to this prolonged requirement for corticosteroid therapy, FDA-approved doses of ruxolitinib (10 mg twice daily) were initiated while corticosteroids were continued at a constant dose. One patient was directly initiated on ruxolitinib since he had mild symptoms, moderate neuroinflammation on brain imaging, and moderate CSF cellular findings and did not favor corticosteroid therapy after a discussion of risks of benefits. Additionally, he had a positive serological test for Chaga’s disease and corticosteroids are not preferred in this setting (22).

At the time of cryptococcal meningoencephalitis diagnosis, all 5 patients received a combination of liposomal amphotericin B and flucytosine for at least 2 weeks followed by fluconazole as antifungal therapy and one of 5 patients had a VP shunt inserted for elevated intracranial pressure. Three out of five patients had been on greater than 6 months of corticosteroids prior to being started on ruxolitinib. Three patients had developed worsening cataracts and osteopenia due to corticosteroids use and one of these patients also developed steroid-induced psychosis, hallucinations and worsening hyperglycemia as a result of prolonged corticosteroids use. One patient (P1) had previously been treated with the IL-6 receptor alpha (IL-6Rα) antagonist tocilizumab(23) with 4 doses (4-8 mg/kg) given over 4 months (23), but was discontinued due to liver enzyme abnormalities. Details of patient presentation, comorbidities, and MRI findings at PIIRS diagnosis are provided in Table 3.

**Table 3.** Demographics and characteristics of patients with post-infectious inflammatory response syndrome.

| No. | Age at diagnosis<br>Race<br>Sex<br>*Cryptococcus* | Baseline<br>CSF CD4<br>counts<br>(cells/uL) | Presenting<br>symptoms | MRI brain findings at PIIRS<br>diagnosis | Co-morbidities | Corticosteroid<br>Treatment<br>Duration<br>(months) | Ruxolitinib<br>Treatment<br>Duration<br>(months) |
| --- | --- | --- | --- | --- | --- | --- | --- |
| P1 | 40-50 year old<br>African American<br>Male<br>*C. gattii* | 414 | Headaches, hearing<br>loss | Hyperintense oval mass in the<br>right superior frontal lobe with<br>eccentric enhancement | Anxiety,<br>Depression,<br>Hypothyroidism,<br>Alcohol use,<br>Hypertension,<br>Osteopenia | 33 | #1: 3<br>#2: 14 |
| P2 | 40-50 year old<br>African American<br>Male<br>*C. neoformans* | 309 | Altered mental<br>status | Leptomeningeal enhancement,<br>communicating<br>hydrocephalus, arachnoiditis | Type II Diabetes<br>mellitus, ischemic<br>heart disease,<br>chronic kidney<br>disease | 19 | 2 |
| P3 | 55-65 year old<br>Caucasian<br>Male<br>Unknown | 561 | Headaches, hearing<br>loss, gait instability | Focal leptomeningeal<br>enhancement within the<br>superior frontal sulcus,<br>enhancement within the<br>internal auditory canals<br>bilaterally | None | 15 | 3 |
| P4 | 20-30 year old<br>Caucasian<br>Male<br>*C. grubii* VNI | 1103 | Headaches, hearing<br>loss, blurred vision | Multiple peri-ventricular and<br>subcortical white matter<br>hyperintense lesions,<br>enhancement in the internal<br>auditory canals | Opioid and<br>marijuana use<br>disorder | 2 | 2 |
| P5 | 40-50 year old<br>Hispanic<br>Male<br>Unknown | 741 | Headaches,<br>Double vision, gait<br>instability | Ventriculomegaly,<br>periventricular edema,<br>communicating<br>hydrocephalus, ependymitis,<br>ventriculitis | Positive serology<br>for South<br>American<br>Trypanosomiasis | None. | 5 |

As shown in Fig. 5, ruxolitinib treatment resulted in reduced CSF markers of inflammation including CSF WBC (Fig. 5a) with a trend towards reductions in total CD45+ CSF cells (Fig. 5b), as well as CSF protein levels (Fig. 5c), but without changes in CSF glucose (Fig. 5d). Flow cytometry of patient CSF samples demonstrated overall reductions in cellular markers of inflammation, including reductions in CSF HLA-DR^+^ CD4^+^ cells (Fig. 5e, f), HLA-DR^+^CD8^+^ cells (Fig. 5g, h), and CD56^+^ NK cells (Fig. 5i, j). A median 4.3-fold reduction of CSF HLA-DR^+^ CD4^+^ cells was observed, which has utility as a treatment biomarker of PIIRS (6). Significant myeloid cell reductions were also observed after ruxolitinib with reductions in overall CSF CD14^+^ monocytes (Fig. Fig. 5k,l) and both innate monocytes (Fig. 5m, n) and mature monocytes (Fig. 5m, o). Pro-inflammatory CSF cytokines and soluble markers were measured (Fig. 5p) and sIL-2R, a soluble marker of T-cell activation, and the cytokine IL-10 were found to be significantly reduced (p < 0.05) following Ruxolitinib treatment. A trend in reductions was noted for the neutrophil chemokine IL-8 (p = 0.09) and pro-inflammatory cytokine IL-6 (p = 0.063) without changes in the Th2 cytokine, IL-13 (p = 0.21).

**Fig 5.**
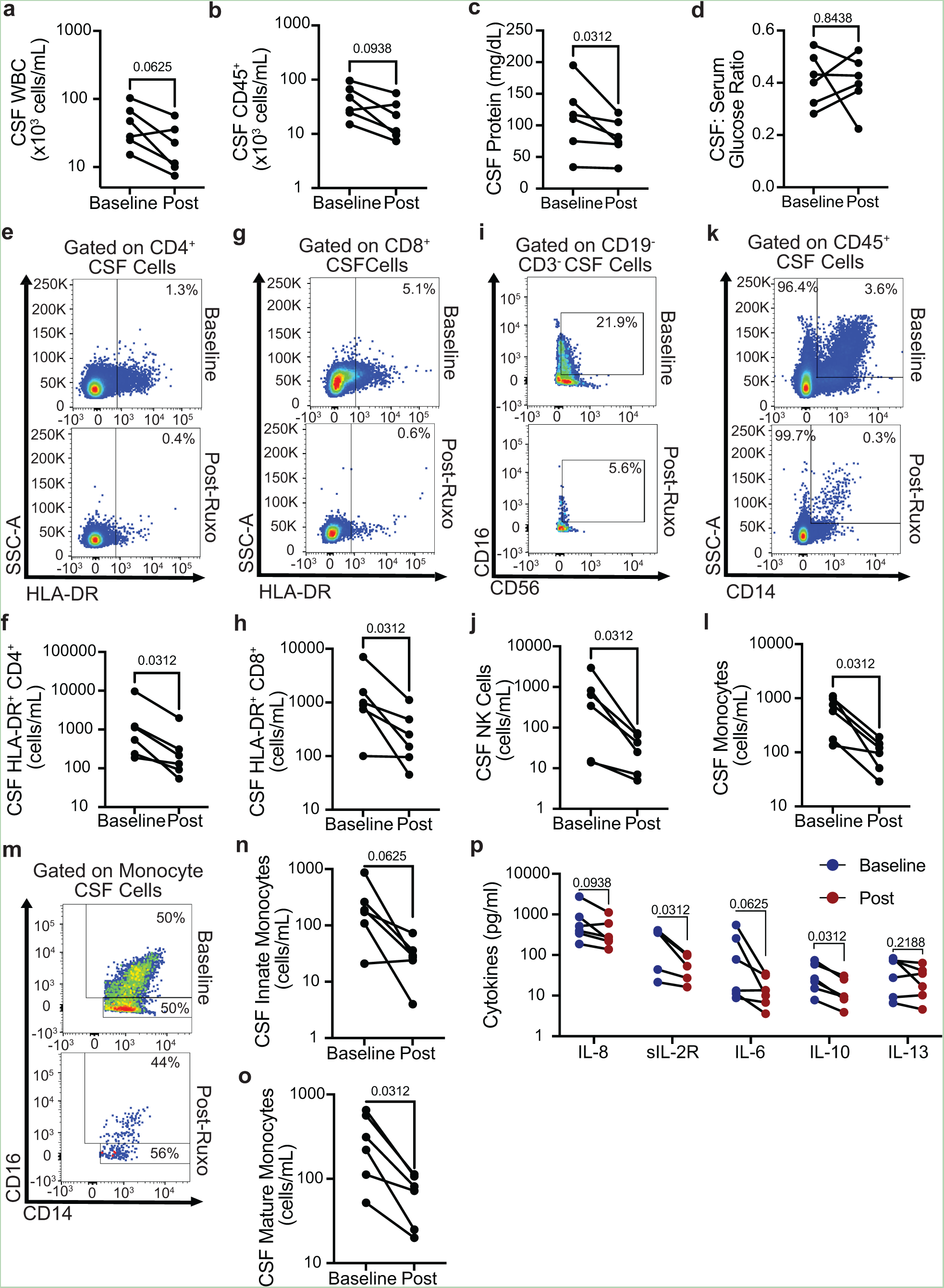
Ruxolitinib reduced inflammatory CD4+ and CD8+ lymphocytes as well as monocytes and soluble markers of CSF inflammation in PIIRS patients. Lumbar punctures were performed before and after 6 1-month courses of ruxolitinib in 5 patients on constant doses of corticosteroids and CSF obtained for testing. **a** WBC counts, **b** CSF CD45+ cells counts, **c** CSF protein, **d** glucose/serum ratio were assessed by commercial clinical testing. **e-o** CSF T cell activation (HLA-DR+ expression) on CD4 (**e,f)** and CD8 T cells (**g, h)**, CD56+ NK cells (**i, j)**, and CD14+ monocytes (**k, l)**, including both innate (**m, n)** and mature monocytes (**m, o)** were measured by flow cytometry at baseline (post-pulse, prior to ruxolitinib treatment) and 1 month following ruxolitinib initiation. Representative and summary data shown. **p** Concentrations of indicated soluble cytokines were measured from the CSF by a commercial assay as described in Methods. Data represents n=5 patients with 6 treatment courses, CSF parameters analyzed using the Wilcoxon-matched pairs signed rank test.

**Fig 6.**
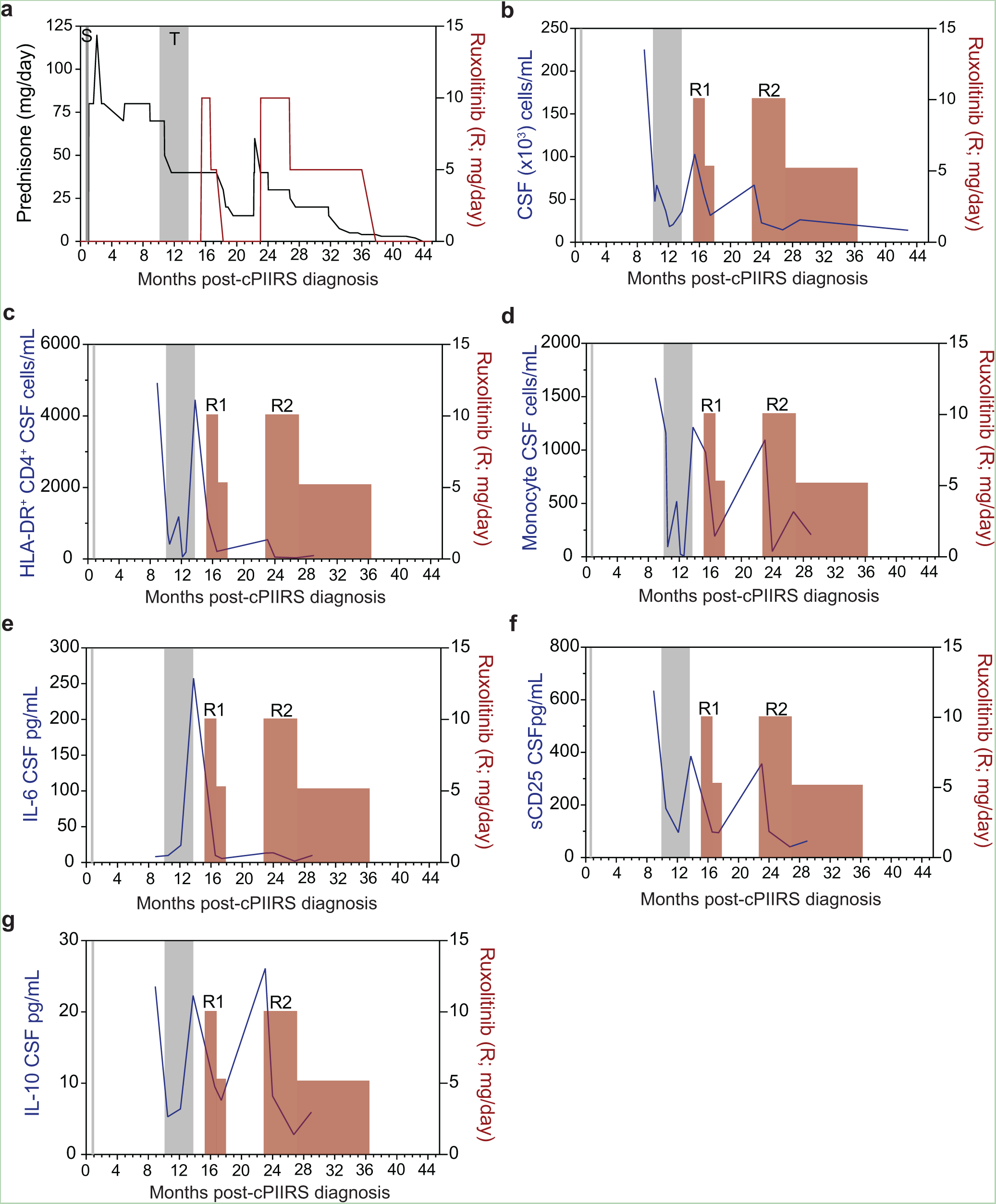
Representative patient treatment timeline (patient 1) and longitudinal cerebrospinal fluid findings. **a** Following c-PIIRS diagnosis, patient was treated with pulse solumedrol (S-vertical gray bar) followed by prednisone taper. Due to a rebound in headaches and mental status changes, the patient required an oral pulse of up to 120 mg of prednisone and a prolonged course of high doses of prednisone. The patient received intravenous infusions of tocilizumab (T-vertical gray bar) while maintained on prednisone until he experienced a rebound in these parameters after a delay in tocilizumab due to LFT abnormalities and tocilizumab was discontinued. Two courses of ruxolitinib (R#1 and R#2-red vertical bar) were initiated with decreases to CSF cell numbers (**b**), HLA-DR^+^ CD4 cells (**c**), monocytes (**d**), IL-6 (**e**), sCD25 (also known as soluble IL-2R) (**f**), and IL-10 (**g**) following each course of ruxolitinib with prednisone finally successfully tapered after the second course.

All patients were monitored with weekly complete blood counts and liver function tests to evaluate acute toxicity. In addition, the patient with a positive serology for Chagas disease underwent EEG and cardiac echo which was normal as well as monthly Chagas PCRs which remained normal. All tolerated, ruxolitinib well without evident toxicity (Table 4) except one patient who developed a 5-fold increase in transaminase, which normalized with a reduction in ruxolitinib and abstinence from an episode of alcohol abuse. For the 4 patients on corticosteroids prior to starting ruxolitinib, prednisone was able to be tapered off while on ruxolitinib although one patient (Patient 2) was treated concurrently at the beginning with a parenteral pulse of corticosteroids before he was eventually tapered off. The mean duration for ruxolitinib used in these patients was 4.8 months (±4.6 months STD; Table 3). Two patients required continued immune suppression and were continued on ruxolitinib for up to 1 year with symptomatic improvements in headaches and concentration sufficient to return to work. All patients were continued on fluconazole while on immunosuppression and no recurrence of CSF fungal growth was noted. Overall clinical benefit may have been evident, but small numbers and lack of placebo controls make such conclusion difficult.

**Table 4.**
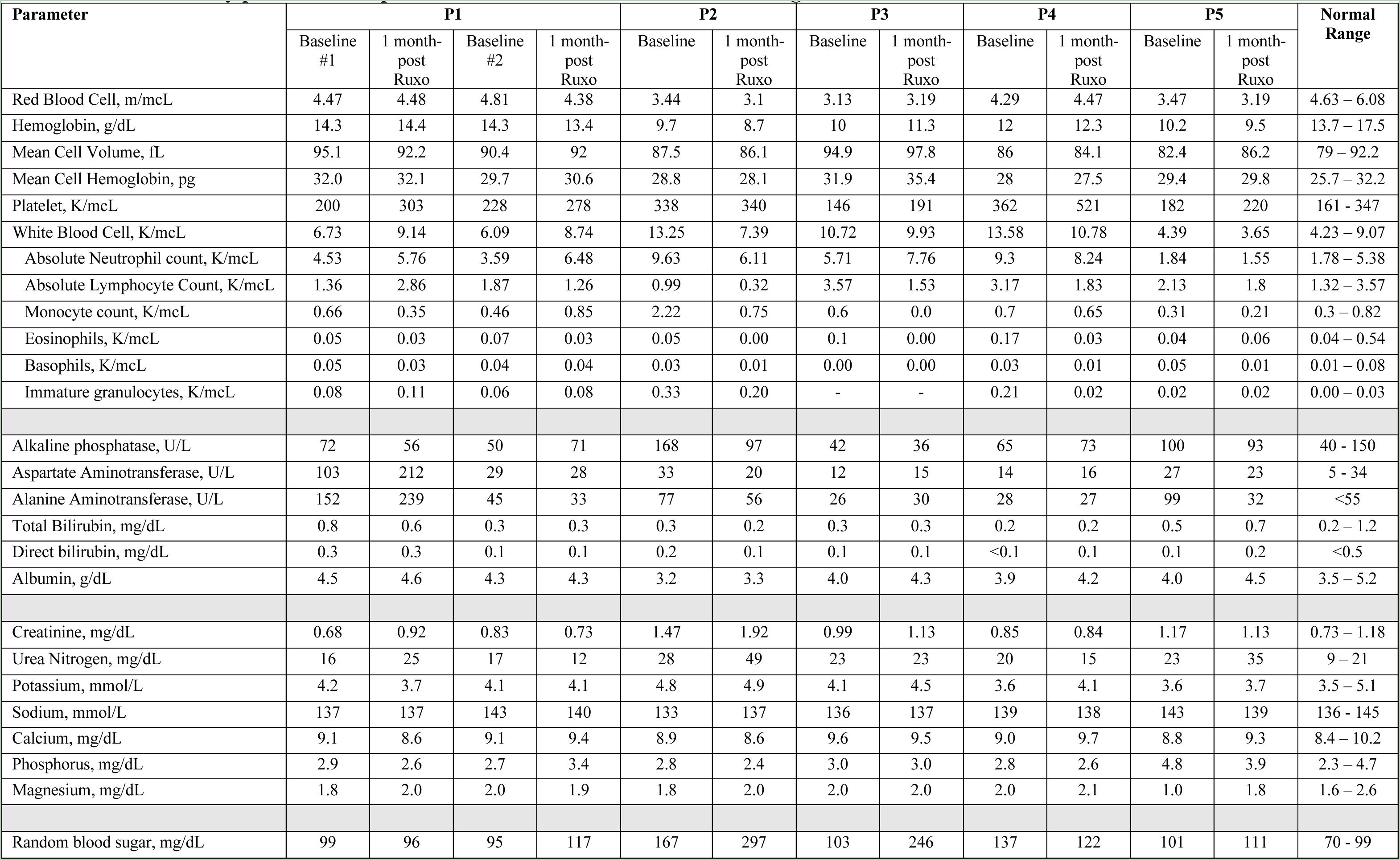
Laboratory parameters of patients before and one month after starting Ruxolitinib treatment.

### Radiological Outcomes

Brain MRI with contrast was performed on all 5 patients at baseline prior to and approximately 1 month after initiation of ruxolitinib therapy. Examples of one patient with PIIRS are shown in Fig. 7a-b with marked decrease in parenchymal and meningeal enhancement after ruxolitinib therapy. Results of aggregate MRI scores defined previously (20) are shown in Figure 7c. In all patients, a reduction in pathologic scores was noted post-ruxolitinib with a significant improvement overall. The most common radiological findings reported at baseline were meningeal enhancement (5 patients), hydrocephalus or shunted ventricular system (5 patients), non enhancing basal ganglia lesions (3 patients), and parenchymal enhancement (2 patients). In patients with hydrocephalus, 1 already had VP shunts in place at the time the baseline MRI was performed. In summary, these data suggest that ruxolitinib in FDA-approved dosages reduced CSF inflammation in this small cohort of cryptococcal PIIRS and was well tolerated, supporting further study as a potential steroid-sparing agent in this disease. The data also suggest that CSF transcriptomics of immune pathology may be useful in the design of innovative therapeutics for inflammatory diseases such as PIIRS.

**Fig 7.**
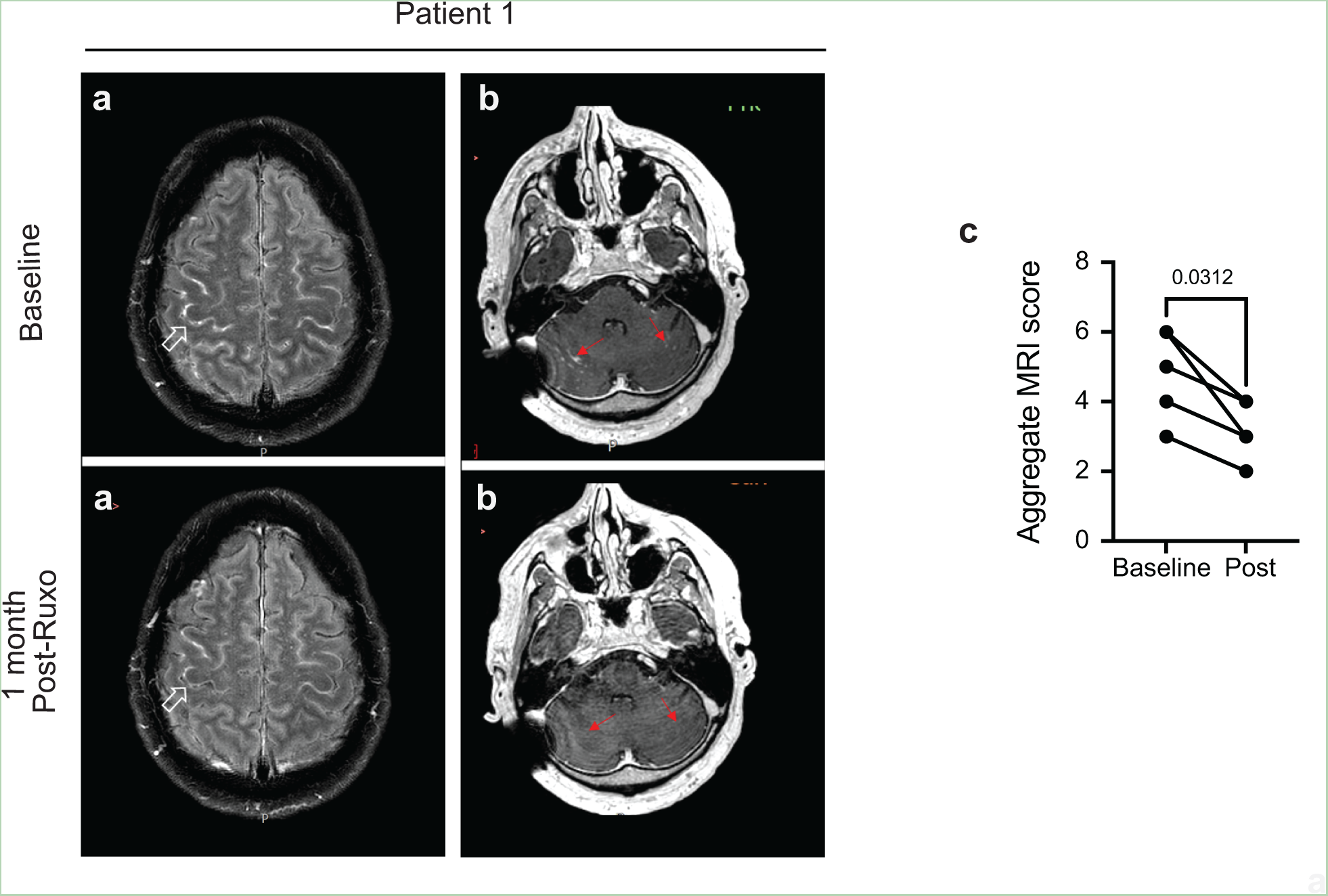
Brain MRI Imaging demonstrates improvement in PIIRS-relate neurological findings in patients after ruxolitinib treatment. **a)** Axial post contrast FLAIR images of the brain. At baseline, meningeal enhancement is observed in multiple convexity sulci. Post ruxolutinib, meningeal enhancement has improved (open white arrow). **b**, Post-contrast T1 weighted images in patient #1 demonstrating abnormal enhancement (solid red arrow) along the cerebellar folia bilaterally which resolved 1 month post-ruxolitinib. **c**, Aggregate MRI scores. Aggregate MRI scores obtained prior to ruxolitinib treatment (baseline) and 1-month post-ruxolitinib treatment initiation (n=5). Wilcoxon-matched pairs signed ranked test All scores decreased on follow-up with p-value indicated above comparison.

## Discussion

JAK/STATs are critical bridges linking cytokines and chemokines extracellularly with transcriptional changes intracellularly resulting in immune activation and inflammation (24). In the context of immune control of pathogens, this signaling pathway is vitally important as humans with variants in JAK and STAT family proteins are highly susceptible to infection by a variety of pathogens (25–29). In response to pathogens and at the molecular level, cytokine stimulation of their cognate receptors leads to JAK kinase activation resulting in the phosphorylation of STATs, which then become activated and translocate as dimers to the nucleus to activate gene transcription. As we have come to appreciate in recent years, these same inflammatory transcriptional programs and pathways necessary for protective responses to pathogens lead to pathology and host damage in the post-infectious period (30) resulting in significant post-acute sequalae exemplified recently by COVID-19, but also present in other syndromes (31) including PIIRS following infections with the fungus *Cryptococcus* including CM. Recently, a parabola has been utilized as an instructional device within a damage-response framework to understand host damage during infections as a function of either microbial factors on one hand or an overly vigorous immune products on the other (1, 30, 32).

Recently, a post-infectious inflammatory response syndrome has been identified in non-HIV-infected individuals with cryptococcal meningitis who have achieved microbiological control with the use of anti-fungal therapies, but deteriorate significantly with high mortality, due to an overly active immune response (5). A hallmark of PIIRS in humans and mouse models is compartmentalized inflammation in the brain with high levels of cytokines in the CSF (6) and brain parenchyma (16), respectively. Neurological infections such as CM are particularly sensitive to inflammatory activation due to the confines of the skull that often results in tissue compression and even brain herniation with high mortality. While corticosteroid therapy was found to provide relief in patients with PIIRS, significant long term side effects may limit the effective use of this agent (6). Thus, the goal of the present study was to utilize a recently-described mouse model of PIIRS (9) to identify predominant pathways involved in PIIRS that could be more specifically targeted by the ever-enlarging armamentarium of biologics. However, targeted therapy directed at the most predominant identified mechanism(s) may reduce the significant sequelae of biologics including fungal infections themselves (33). For example, identifying predominant underlying mechanisms responsible for exuberant inflammation and host damage in similar immune mediated diseases as resulted in the safety of several JAK inhibitors that have now gained FDA-approval for conditions such as COVID-19 (34), ulcerative colitis (35), and graft-versus-host disease (36).

Thus, to determine the causal pathway responsible for the exuberant inflammation in PIIRS and potential therapeutic target(s), we analyzed transcriptional data from the brains of PIIRS model mice to rank the top transcriptional regulators of PIIRS-associated neuroinflammation and identified JAK/STAT signaling as predominant by neuro-inflammation-related transcripts. This suggests similarities with inflammatory disorders such as rheumatoid arthritis (RA) where JAK/STAT pathways are important mediators of disease (37). This has led to the use of inhibitors such as ruxolitinib being utilized for this disease, as well as others such as systemic lupus erythematosus and vasculitides (38). However, it is important to realize that multiple inflammatory pathways are likely to play a role, similar to RA that also have roles for the NF-κB pathways (39) and IL-6 (40), that latter of which was found to be less predominant in the present studies but has shown some effectiveness in PIIRS (23). However, in the present study, the orally available ruxolitinib was more convenient and did not expose patients to infusion-related toxicities from tocilizumab and resulted in greater degrees of CNS inflammatory suppression than tocilizumab in standard doses. Nevertheless, the inflammatory transcription profile of PIIRS appeared to be distinguished from other syndromes, such as C3-glomeruolopathy that appear to be more predominantly complement-mediated syndromes (41, 42). Inflammasome activation pathways also did not appear to be as predominant, distinguishing PIIRS from syndromes such as cryopyrin-associated periodic syndromes or various non-infectious neurodegenerative diseases (43). Based on this data, we hypothesized that JAK inhibition may diminish neuroinflammation and lessen disease. Indeed, using 2 murine models, we showed that the JAK1/2 inhibitor, ruxolitinib, significantly reduced the manifestation of PIIRS in mice. Specifically, through reducing systemic immune activation/proliferation (splenomegaly), cerebral edema, CNS innate and adaptive immune cell accumulation and activation (particularly effector CD4 T cells and inflammatory myeloid cells), neuropathology, and transcriptional signatures of adaptive immune cytotoxicity and cytokine signaling. We did find that treatment of mice without anti-fungal therapy resulted in an increased fungal burden, and concomitant antifungal therapy reduced this and led to improvements in weights, a major surrogate for improvement in this model (9).

This encouraging pre-clinical data then led to the use of ruxolitinib in standard doses for 5 patients who had continued inflammation and slow clinical response despite corticosteroid therapy. Importantly, in a small cohort of PIIRS patients, ruxolitinib treatment while maintaining a constant corticosteroid dose resulted in clinical improvement with reduced biomarkers of inflammation in the CSF. Retrospective analysis of this data found significant reductions in inflammatory parameters, including a cellular marker, CSF HLADR^+^CD4^+^ cells, used previously as a surrogate of treatment response (6). Patients subjectively noted subjective clinical improvements with two patients being able to return to work who were previously unable; however, the small number and lack of controls prevents any conclusions of ruxolitinib efficacy in PIIRS.

Limitations to this study: This is a small cohort study and not sufficiently powered or controlled for to determine efficacy of ruxolitinib for treatment of PIIRS. Thus, this study is also underpowered to make definitive conclusions of immunosuppressive complications or toxicities. One patient developed elevations in liver enzymes but was found to relapsed alcohol abuse which improved during abstinence and a reduction of dose or ruxolitinib. Further randomized studies will be needed to determine if the clinical efficacy of steroids can be potentiated by ruxolitinib and/or whether it may benefit long-term outcomes and reduce corticosteroid requirements in PIIRS. Additionally, though NanoString analysis of 757 genes revealed a significant and causal role for JAK/STAT-signaling in mediating neuroinflammation and host damage in PIIRS, a more comprehensive transcriptional analysis utilizing single cell-RNA sequencing of patient CSF and mouse brain samples may reveal additional therapeutic targets for the treatment of PIIRS.

In summary, this is the first report identifying a predominant role for JAK/STAT signaling in a fungal inflammatory disease. Furthermore, pathway-directed therapeutics utilized for the first time ruxolitinib treatment for steroid-refractory PIIRS, in which 5 patients had measured and meaningful reductions in biomarkers of CNS inflammation while maintaining a constant dose of corticosteroids. Importantly, all patients were able to taper completely off of corticosteroids and at this time all patients are clinically stable with continued negative fungal cultures after completion of immunosuppressant therapy.

## Methods

### Ethics statement

The National Institute of Allergy and Infectious Diseases (NIAID) Institutional Review Board (IRB) approved this study under NIAID Protocol number 93-I-0106. All subjects provided written informed consent directly or via their durable power of attorney.

### Study Design and Participants

All five patients were seen at the National Institutes of Health (NIH) Clinical Center, Bethesda, MD between 2020 and 2023 as part of a prospective observational study examining host genetics and immunology of cryptococcal disease in previously healthy, non–HIV-infected adults. Patients were excluded if they were <18 years old and had any underlying immune deficiency, such as HIV or receipt of immunosuppressant medications, including cancer chemotherapy or monoclonal antibodies. A diagnosis of CM was defined as a positive latex agglutination cryptococcal antigen or the isolation of *Cryptococcus* in one or more CSF cultures, or both. The five patients analyzed for the present study had completed standard therapy of liposomal amphotericin B and flucytosine at another institution prior to NIH transfer and had negative CSF fungal cultures. All patients underwent referral to the NIH after they developed refractory symptoms consistent with PIIRS, including declining mental status and cranial nerve defects, and subsequently met criteria for PIIRS.

### Protocol

At the NIH, all patients were diagnosed with PIIRS (6) and 4 patients (P1-P4) received pulse methylprednisolone 1 gram parenteral daily for 7 days followed by 1 mg/kg/day prednisone for 1 month followed by a taper as tolerated. Lumbar punctures were performed on admission, after pulse methylprednisolone, and as clinically required based on opening pressures and symptoms. Ruxolitinib was utilized in patients poorly responsive to corticosteroid therapy and data was collected retrospectively. Spinal fluid was sent for routine analysis, fungal cultures, cryptococcal antigen titers, immunophenotyping, soluble cytokine analysis as previously described (5, 44) and CSF fluid component (glucose and protein) measurements. Retrospective analyses of CSF studies were reviewed to generate data for these studies. Neuroimaging of the brain via MRI was performed throughout treatment as previously described (45).

Radiological findings were scored based on the presence of 9 criteria; details of scoring can be found in (6). Aggregate scores were calculated for each patient ranging from a minimum of 0 to a maximum of 12. A decrease in aggregate scores on post-ruxolitinib MRI compared to baseline indicated radiological improvement.

### CSF and PBMC collection, processing, and immunophenotyping by flow cytometry

CSF was collected on ice and processed immediately. A portion of the collected CSF (10–20 mL) was collected and assigned prospective alphanumeric codes. To determine whether viable or viable but not culturable (VBNC) *Cryptococcus* cells were present in patient CSF samples, 100 μL CSF was plated on YPD media or Minimal Media containing 125 μM pantothenic acid (reactivation media)(46, 47), respectively, and grown at 37°C, 5% CO_2_ for at least 1 month. No patient CSF samples grew *Cryptococcus* on these media at any time throughout their treatment at the NIH. CSF volume and visual appearance were recorded, and CSF samples were centrifuged at 300 × *g* for 10 min at 4°C within 15 min of collection. The CSF supernatant was aliquoted and stored in polypropylene tubes at −80°C and was tested for soluble cytokines by ARUP Laboratories, Salt Lake City, UT, by a method previously described (5). Cell pellets were resuspended in 500 uL PBS containing 1% BSA and counted using a Luna-fl Dual Fluorescence Cell Counter (Logos Biosystems), and the number of white blood cells (WBCs) per mL of CSF was calculated. A minimum of 5 × 10^4^ viable CSF cells were analyzed immediately by 9-color flow cytometry to enumerate absolute numbers of 10 subsets of CSF immune cells. A minimum of 2 x10^5^ CSF cells were spun down and resuspended in 1 mL of RNAlater (Millipore Sigma, R0901) and stored at -80C for NanoString analysis.

Peripheral blood mononuclear cells (PBMC) isolation from whole blood was performed by density gradient centrifugation using Ficoll-Paque PLUS Premium (GE Healthcare, 17-5442-03) and SEPMate-50 tubes (STEMCell, 85450).

To assay for immune cell populations within the CSF and peripheral blood, CSF cells and PBMC were first blocked with Fc Receptor Binding Inhibitor (Invitrogen) in flow staining buffer (PBS containing 1% BSA) for 15 min at 4°C and then stained with antibodies to CD45 FITC (HI30, eBiosciences), CD3 PB (UCHT1, Biolegend), CD4 PE-Cy7 (SK3, BD Pharmingen), CD8 APC (SK1, Biolegend), HLA-DR eFluor605NC (LN3, eBiosciences), CD19 AlexaFluor700 (HIB19, Biolegend), CD59 (NCAM) PE/Dazzle594 (HCD56, Biolegend), CD14 PerCP-Cy5.5 (63D3, Biolegend), and CD16 APC-H7 (3G8, BD Pharmingen) for 30 min at 4°C. After incubation, cells were washed twice with flow staining buffer and fixed in Cytofix buffer (BD) for 30 min prior to washing. Data was immediately collected on a LSRFortessa flow cytometer (BD) with FACSDIVA Software (BD) and analyzed using FlowJo software (BD). Appropriate unstained, single color control and fluorescence-minus one (FMO) controls were run with samples.

All events were first gated using Time versus FSC-H to exclude events at the start and end of the run, which often have altered light scattering profiles. Next, debris and cell aggregates were excluded using FSC-H versus FSC-W (gated for FSC singlets) and SSC-H versus SSC-W (gated for SSC singlets) followed by gating on CD45+ CSF cells (SSC-H versus CD45) to exclude non-leukocytes. Activated CD4+ T cells were defined as CD45^+^CD14^−^CD3^+^CD19^−^CD4^+^HLA-DR^+^. Activated CD8+ T cells were defined as CD45^+^CD14^−^CD3^+^CD19^−^CD8^+^HLA-DR^+^. Classical (mature) monocytes were defined as CD45^+^CD14^+^CD16^−^HLA-DR^+/−^(48). Non-classical (innate) monocytes were defined as CD45^+^CD14^+^CD16^+^HLA-DR^+/−^(48). Unstained and FMO control samples were used to define negative and positive populations for gating.

### Mice

All animal experiments were conducted under protocols approved by the Institutional Animal Care Committee (IACUC) of the Intramural NIH/NIAID and were performed in accordance with NIH guidelines and the *Guide for the Care and Use of Laboratory Animals* under protocol LCIM 12E (PHS assurance #: D16-00602). C57BL/6 female mice were obtained from Taconic Biosciences, Inc. (B6NTac, Germantown, NY). Mice were housed under specific pathogen free (SPF) conditions in microisolator cages at the NIH and were provided with food and water ad libitum. Mice were 6 to 12 weeks old at the time of infection and were humanely euthanized by isofluorane inhalation at the time of data collection. For mortality studies, mice were euthanized when they lost 20% body weight, had persistent cranial swelling, and/or developed neurological symptoms.

### Ruxolitinib treatment

Ruxolitinib (R-6688, LC Laboratories) was formulated into Nutra-gel diet (F5769-Kit, Bio-Serv) at a concentration of 1 g/kg. Mice were acclimated to untreated Nutra-gel diet (replaced daily) alongside normal rodent chow for 7 days. Mice were provided with Ruxolitinib diet or Nutra-gel diet ad libitum (replaced daily) for 2 days prior to infection and continued throughout infection study period. In experiments where mice were treated with Amphotericin B during infection, Ruxolitinib diet was added on day 18 post-infection and continued throughout the study period.

### Amphotericin B treatment

Amphotericin B (AmB, Sigma A9528) was dissolved in sterile pharmaceutical-grade saline at 1 mg/mL. Mice received a daily dose of 5 mg/kg AmpB beginning on day 14 post-infection, which continued throughout the study period.

### C. neoformans and PIIRS model mouse infection

ATCC 24067 (American Type Culture Collection, Manassas, VA), *C. neoformans* 52D strain, was used to infect mice in this study. Cryptococcal cells from frozen stocks (10% glycerol) were plated on Yeast Peptone Dextrose (YPD, BD Difco) agar plates containing 50 mg/ml chloramphenicol (Sigma) at 30°C. Single colonies were subsequently picked and grown overnight in YPD broth (BD Difco) containing 50 mg/ml chloramphenicol at 30°C with 225 rpm shaking. Fungal cells were washed three times in PBS, counted on a hemocytometer with trypan blue, and adjusted to a concentration of 5 × 10^6^ per mL before infection. Mice were infected with 10^6^ yeast (in 200 μl of sterile PBS) via tail-vein intravenous injection. Serial dilutions of the *C. neoformans* suspension were plated on YPD agar to confirm the number of viable fungi in the inoculum. Weights were monitored three times a week. For analysis of brain fungal burdens, animals were euthanized and organs weighed, brains homogenized in PBS, and serially diluted before plating onto YPD agar supplemented with chloramphenicol (Sigma). Colonies were counted after incubation at 30 °C for 48 hours. For analysis of brain cytokines, brain homogenates were diluted 2-fold in PBS before measurement using the Mouse Cytokine 32-plex Discovery Assay (Mouse Cytokine Array/Chemokine Array 32-Plex Panel; Cat#: MD31; Eve Technologies, Alberta, Canada).

### Analysis of murine cells by flow cytometry

Leukocytes in the brain were isolated as previously described (9). Briefly, mice were euthanized and perfused with 10 ml of PBS to remove circulating red blood cells and leukocytes from the brain. The brains were aseptically removed, transferred to gentleMACS C tubes containing 5 ml of sterile complete RPMI 1640 (with 5% fetal bovine serum, 25 mM Hepes, GlutaMAX, penicillin-streptomycin, and enzyme cocktail containing collagenase (50 ug/mL; Roche), hyaluronidase (100 U/mL; Roche), and DNase (100 U/mL; Roche)). The tissue was processed on a gentleMACS homogenizer (Miltenyi Biotec) and incubated at 37°C for 45 minutes. Small samples of the homogenate were collected immediately after processing for whole-brain fungal burden, RNA, and cytokine measurements. The remaining homogenate was washed with RPMI 1640 and filtered through a 70-μm cell strainer. A 33%/67% Percoll (GE Healthcare)/RPMI media gradient was used to remove cell debris, myelin, and neurons, and then microglia and brain-infiltrating leukocytes were recovered from the pellet. Suspensions were centrifuged at 8000 g for 30 min at 4 °C with the brake off. Isolated cells were washed twice with RPMI 1640 (with 5% fetal bovine serum, penicillin-streptomycin) to remove residual Percoll before use in assays. Total cell counts were obtained by counting live cells using a Luna cell counter.

For splenocyte isolation, spleens were removed and then mechanically dispersed by using a 3-ml sterile syringe plunger to press through a 70-μm cell strainer (BD Falcon, Bedford, MA) in complete RPMI 1640 medium. The cell suspension was washed with PBS and centrifuged. Erythrocytes in the cell pellets were lysed by the addition of 5 ml of NH_4_Cl buffer [0.829% NH_4_Cl, 0.1% KHCO3, and 0.0372% Na_2_EDTA (pH 7.4)] for 5 min, followed by addition of a 10-fold excess of RPMI 1640 medium. After centrifugation, the splenocytes were saved for further use.

### Flow cytometry

Cells were stained with fixable LIVE/DEAD dye (Life Technologies), blocked with anti-CD16/32, and stained with antibodies for CD45 (30-F11; BioLegend), CD3 (17A2; BioLegend), CD4 (GK1.2; eBiosciences), CD11b (M1/70; BioLegend), CD11c (N418), CXCR3 (CXCR3-173), Ly6C (HK1.4; BioLegend), CD44 (IM7; BioLegend), CD62L (MEL-14; Invitrogen), and/or major histocompatibility complex class II (M5/114.15.2; eBiosciences). For IFN-γ production, the cells were stimulated for 6 hours with phorbol myristate acetate and ionomycin in the presence of brefeldin A and monensin for the final 4 hours. The cells were stained for extracellular markers and then fixed with fixation/permeabilization buffer, and intracellular staining for Foxp3 and IFN-γ was performed in permeabilization buffer. Fluorescence minus one controls, single color controls, and an unstained control were used for all experiments. Data were collected on an LSRFortessa (BD) and were analyzed using FlowJo (BD). Microglia are defined as CD45intCD11b+, while inflammatory monocytes are CD45hiCD11b+Ly6c+.

### NanoString analysis

Total RNA was isolated from the whole-brain homogenate using TRIzol (Life Technologies) and gene expression analyzed using the NanoString nCounter Mouse Neuroinflammation Panel (NanoString Technologies). Total RNA was isolated from human patient CSF cells stored in RNAlater using buffer RLT and the RNeasy Micro Kit (Qiagen) and gene expression analyzed using the NanoString nCounter Human Neuroinflammation Panel (NanoString Technologies). Data analysis was performed on the nSolver analysis software according to the manufacturer’s instructions and built-in statistical analyses. Pathway scores were calculated as the first principal component of the pathway genes’ normalized expression. The software will orient the scores such that increased score corresponds with increased expression in a majority of the pathway genes. Scores of the cell type characteristic genes were analyzed using the default settings from the cell type profiling algorithm in the nSolver analysis software.

### Western Blot Analysis

Western blotting using indicated anti-mouse target antibodies was performed. For western blotting, brains were homogenized in Pierce RIPA Buffer (Thermo Fisher Scientific, 89900) with the addition of Halt Protease and Phosphatase Inhibitor Single-Use Cocktail (100x) (Thermo Fisher Scientific, 78442) and protein concentrations were determined using a colorimetric assay (Bio-Rad, 5000006). 80 micrograms of each sample were subjected to a 12% Mini-PROTEAN® TGX™ Precast Protein Gels, 10-well, 50 µl (Bio Rad, 4561043) and proteins were transferred onto nitrocellulose membranes (GenScript, L00224). Membranes were immunoblotted with rabbit anti-phospho-Stat1 (Tyr701) (Cell Signaling, 58D6, 9167), rabbit anti-Stat1 (Cell Signaling, D1K9Y, 14994), rabbit anti-phospho-Stat3 (Tyr705) (Cell Signaling, D3A7, 9145), rabbit anti-Stat3 (Cell Signaling, 79D7, 4904), and rabbit anti-beta-actin (Cell Signaling, 13E5, 4970). Membranes were then probed with Goat anti-Rabbit IgG (H+L) Cross-Adsorbed Secondary Antibody, HRP (Thermo Fisher Scientific, G-21234) and proteins were visualized using Clarity Western ECL Substrate (Bio-Rad, 170-5061).

### Histopathology

Mouse brains were removed at indicated time points and fixed in 10% formalin for 24 hours before embedding in paraffin and sectioning. Tissue sections were stained with hematoxylin and eosin and Gomori’s methenamine silver. Sections were analyzed by MoticEasyScan Pro6 and Motic Digital Slide Assistant software (Motic).

### Fluorescent Microscopy

The brains were fixed with 4% PFA, cryoprotected in 30% sucrose for 72 hours, then frozen and embedded in OCT compound. Antigen retrieval was carried out in citrate buffer in a boiling water bath for 15 min. Sections were blocked in 5% bovine serum albumin (BSA) in PBS. For the beta III tubulin and synaptotagmin-7 staining, sections were stained with a rabbit polyclonal beta III antibody (1:100, Abcam) and synaptotagmin-7 antibody (clone S275, Invitrogen) for 1 h at 37°C. Secondary antibody staining was done with goat anti-mouse Alexa 555 antibody and goat anti-rabbit Alexa 488 antibody (1:500, Invitrogen) for 45 min at 37°C. For the beta III tubulin and cleaved caspase-3 staining, sections were then stained with a primary antibody to beta III tubulin (TuJ1), a rabbit antibody specific to cleaved caspase-3 (1:100, Sigma), and a rat anti-mouse CD45 (clone 30-F11 from Biolegend) for 1 h at 37°C. Sections were then washed in PBS and counterstained with fluorophore-conjugated a secondary antibodies: goat anti-rabbit Alexa 555, goat anti-mouse Alexa 488, and donkey anti-rat Alexa 647 antibody (1:500) for 45 min at 37°C. After that, the sections were washed in PBS for 30 min. The auto fluorescence was quenched by Vector’s true view kit (Vector Laboratories) as per the manufacturer’s protocol before mounting with a mounting medium. Samples were visualized using a KEYENCE microscope BZ-X800 series. Images were captured by the camera provided by the vendor by using a ×20 objective 0.45 numerical aperture (NA) Nikon. Images were de-convoluted for enhancing the clarity of the images. All the images were captured under the same exposure conditions. Grayscale mages were pseudo-colored for presentation.

We use linear regression models fit via generalized estimating equations (GEE) to assess the effect of RUXO, with and without AMB, on each of the outcomes. In each of the GEE models, we used an exchangeable working correlation structure and clustered on mouse ID. Lesion diameter and MFI measurements were modeled on the natural log scale with an identity link, hence the exponentiated model coefficients are interpreted in terms of multiplicative changes on the geometric mean outcome. CD45 counts were modeled using a Poisson GEE with loglink. Models were fit using the geepack package in R (49). Given the exploratory nature of this study, no adjustment for multiple comparisons is made.

### Statistical analysis

Statistical analysis was performed using GraphPad Prism version 8 software with unpaired Student’s t test or Wilcoxon-matched pairs signed rank test.

## Data Availability

All data produced in the present study are available upon reasonable request to the authors.

